# Detection and Prevalence of Syphilis, Hepatitis C Virus and *Helicobacter pylori* Co-Infection among Pregnant Women attending Primary Health Centre, Mbodo-Aluu, Rivers State, Nigeria

**DOI:** 10.1101/2024.03.22.24304749

**Authors:** C. C. Adim, F. H. Nnamdi, C. D. Ugboma, T. I. Cookey, H. C. Innocent-Adiele, E.N. Onu, M. U. Igwe, A. M. Awanye, B. J. Okonko, C. M. Enemchukwu, G. A. Nwankwo, I. O. Okonko

## Abstract

Due to the possible negative effects on both the mother and the foetus, co-infection with syphilis, hepatitis C virus (HCV), and *Helicobacter pylori* (*H. pylori*) in pregnant women is a serious public health problem. In this study, pregnant patients at the Mbodo Health Centre in Aluu, Port Harcourt, Rivers State, were asked to rate their prevalence of co-infection and related risk factors. 96 willing pregnant women gave consent for blood samples to be drawn, and ELISA and fast diagnostic kits were used to test the sera for *Helicobacter pylori*, hepatitis C, and syphilis. Results showed that 31.0% of the pregnant women had *Helicobacter pylori*, but none had positive results for syphilis or the hepatitis C virus. The highest prevalence of *H. pylori* infection, as determined by ELISA, was observed in the age group 30-39 years (37.5%), married groups (32.7%), secondary school education (44.0%), students (40.0%), and Christian religion (31.3%). This finding demonstrated that *H. pylori* was more common in the study area than the other two pathogens. Pregnant women visiting the Primary Health Centre, Mbodo-Aluu, Rivers State had significant rates of *H. pylori* infection with no coinfection with HCV and syphilis. This study emphasises the necessity of integrated screening and treatment initiatives during antenatal care. However, preventing unfavourable pregnancy outcomes and lowering the risk of vertical transmission to the baby need the early detection and treatment of these pathogens. Therefore, it is important to emphasise good knowledge and education about the infection in this area.

## 1. Introduction

The spirochete *Treponema pallidum* is the source of the systemic illness syphilis (Norwitz & Hicks, 2024). According to Eppels et al. (2022), syphilis is a treponemal illness that can be contracted sexually, hematogenously, or vertically from mother to child. Even though there are evidence-based curative treatment options and penicillin has been available for more than 70 years, there is still a public health risk associated with it, and its prevalence has been rising recently (Eppes et al., 2022). The possibility of infection spreading to the fetus through the placenta makes it more concerning during pregnancy (Norwitz & Hicks, 2024).

A baby born with congenital syphilis may suffer significant health consequences, however, the degree of the newborn’s health impact will vary depending on when the mother contracted the infection and whether or not she received treatment (CDC, 2020, 2022). Pregnant women with syphilis may experience miscarriage, stillbirth, or the baby’s death soon after birth (CDC, 2020, 2022). 40% of newborns born to women with untreated syphilis may die from the illness or have a stillbirth (CDC, 2020, 2022).

According to Husen and Tadesse (2019), research on the prevalence of syphilis in sub-Saharan Africa is incredibly inconsistent and conflicting. In Cameroon (Tagny et al., 2009, 2010), the prevalence of syphilis was estimated to be 9.1%; 7.9% in Ghana (Adjei et al., 2003); in Nigeria, it was estimated to be 19.3% in Abuja (Sule et al., 2010); in Ibadan, it was 0.0%, 0.8%, 1.5% and 0.0%, respectively (Okonko et al., 2012a, b, c, 2013); 1.5% and 6.6% in Port Harcourt (Adewuyi-Oseni et al., 2019; Okonko et al., 2020a); and in Uyo, it was 1.7% (Okonko et al., 2020b).

The hepatitis C virus (HCV) causes hepatitis C, a liver illness that should be screened for by prenatal care providers for all expectant patients which started in April 2020 (CDC, 2021). Most persons with a chronic HCV infection do not exhibit symptoms, although it can cause liver cancer and cirrhosis (CDC, 2021). Maternal HCV infection has been associated in several studies with unfavourable pregnancy outcomes. A mother can pass on hepatitis C to her child (CDC, 2021). When hepatitis C is detected in pregnant women, treatment can be accessed, and at-risk babies can be identified for further testing and monitoring as needed (CDC, 2021).

New infection rates with HCV rose by almost 60% between 2015 and 2019. Furthermore, adults between the ages of 20 and 39 accounted for about 63% of new HCV infections in 2019 (CDC, 2021). Among those who had live babies, the rate of HCV infection almost doubled between 2009 and 2014 (CDC, 2021). An estimated 29,000 HCV-positive women gave birth annually between 2011 and 2014. Both during pregnancy and childbirth, an infected mother can pass HCV to her unborn child (CDC, 2021). If the nipples are not damaged and bleeding, breastfeeding is not thought to be a means of transmission (CDC, 2021). In 5.8% of pregnancies, moms with HCV infection pass on their infection to their unborn child; this risk increases if the mother also has HIV (CDC, 2021). The CDC expects that the number of perinatal hepatitis C cases found and reported to the CDC will rise as capacity for viral hepatitis surveillance improves (CDC, 2021).

The literature has been acknowledging for a few years now that *Helicobacter pylori* is linked to several extra-gastric disorders. One of the most prevalent chronic infections globally is *Helicobacter pylori*, with prevalence rates ranging from 35 per cent in high-income nations (Zamani et al., 2018; Mesfun et al., 2022) to 79–90% in low-income nations (Asrat et al., 2004a,b; Hooi et al., 2017; Mesfun et al., 2022). Most of the time, the infection is asymptomatic, but it can also cause non-malignant problems including iron-deficiency anaemia, dyspeptic illness, and malignant complications like gastric and oesophagal cancer (Xie et al., 2013; Mesfun et al., 2022). Even during pregnancy, research is being done on how this pathogen affects intestinal issues. In particular, hyperemesis gravidarum, a severe form of morning sickness and vomiting during pregnancy, seems to be associated with this Gram-negative bacterium. The primary debate point is the link between *H. pylori* infection and various ailments (Koa et al., 2016).

The majority of infections are picked up in early life, with Asia and Africa having the highest prevalence (Emerenini et al., 2021). Antibodies against *H. pylori* are developed by infected persons and can last for up to six months following eradication (Emerenini et al., 2021). In our environment, low socioeconomic position, inadequate personal hygiene, poor sanitation, and limited access to potable home water are commonplace and have been associated with a higher incidence of the disease and its associated health implications (Emerenini et al., 2021).

Coinfection is a challenge because it affects the rate at which the disease progresses (Okonko et al., 2020c). If left untreated, syphilis, the hepatitis C virus, and *Helicobacter pylori* can all affect expectant mothers and result in pregnancy-related problems. The frequency of *Helicobacter pylori*, hepatitis C virus, and syphilis have been linked to an increased risk of morbidity and mortality. Given the high incidence of *H. pylori* in Nigeria, it is acceptable to need an effective initial diagnosis, medication, or monitoring of the eradication process (Smith et al., 2019a, 2022a; Bordin et al., 2021; Ahaotu et al., 2023a). Finding out how common co-infections of *Helicobacter pylori*, hepatitis C virus, and syphilis are in pregnant women is the primary goal of this study.

## 2. Materials and Methods

### 2.1. Study Area and design

The study design employed in this work was a cross-sectional hospital survey. It examines the prevalence of *Helicobacter pylori*, hepatitis C virus, and syphilis in the serum of expectant patients visiting the Primary Health Centre, Mbodo-Aluu. Mbodo-Aluu is situated in the Ikwerre local government area of Rivers State, Nigeria. Aluu lays 4.9339^0^N and 6.9437^0^E in Rivers State, Nigeria. Pregnant women who visited Mbodo Health Centre comprised this target demographic.

### 2.2. Study population

In order to ensure that every member of the research population has an equal chance of being picked, a cross-sectional survey technique based in a hospital was used in this work. The aforementioned methodology was employed until a hundred expectant mothers, aged 20-49, and underwent testing.

### 2.3. Sample collection, transport, preparation, and storage

Blood was aseptically extracted into a sterile EDTA tube after the enrolled subject was venepuncture using a 2 ml sterile syringe. Following that, the blood samples were sent directly in an ice pack or cold chain to the Virus & Genomics Research Unit of the Department of Microbiology at the University of Port Harcourt. The plasma was aspirated into sterile Eppendorf tubes using a disposable hand pipette after the blood samples were centrifuged to separate the serum from the red blood cells. The samples for haemolysis were thrown away after being coded to avoid getting erroneous findings. Before being utilised, the blood serum was stored at -20°C.

### 2.4. Serological analysis

Using commercially available ELISA kits (manufactured by DIA.PRO in Milano, Italy), plasma samples were analyzed for *Helicobacter pylori*, Syphilis (*Treponema pallidum*), and HCV under the manufacturer’s instructions.

### 2.5. Data analysis

Data analysis was done using Microsoft Excel. Seropositivity was calculated as the difference, multiplied by 100, between the number of samples that tested positive and the number of samples that were screened. The Chi-square test was used to ascertain the relationships between the sociodemographic characteristics of the participants. P <

0.05 was set as the threshold for statistical significance.

## 3. Results

### 3.1. Characteristics of the pregnant women

This study had 100 pregnant women in total. Following stratification, Table 1 displays the socio-demographic attributes of these pregnant women. Age of the participants ranged from 20 to 49 years old. The age group 30–39 made up 48.0% of the population, followed by the 20–29 (45.0%), and 40–49 (7.0%). Just 49.0% of the study’s participants were married pregnant women, compared to roughly 51.0% of the unmarried (single) participants. Twenty-five per cent of them had completed secondary school, 52 per cent had completed post-secondary education, and 23 per cent had never attended any kind of school. In terms of occupation, students constituted 25.0% of the groups, while non-students made up 75.0% of the groups. All of them identified as Christians (Table 1).

**Table 1:** The Prevalence of Syphilis, HCV, and *H. pylori* with the Socio-Demographic Characteristics of Pregnant Women.

| **Variables** | **No. Tested (%)** | **No. Positive for *H. pylori* (%)** |
| --- | --- | --- |
| **Age Group (years)** |  |  |
| 20-29 | 45 | 12(26.7) |
| 30-39 | 48 | 18(37.5) |
| 40-49 | 7 | 1(14.3) |
| **Marital Status** |  |  |
| Single | 51 | 15(29.4) |
| Married | 49 | 16(32.7) |
| **Educational Level** |  |  |
| Tertiary | 52 | 17(32.7) |
| Secondary | 25 | 11(44.0) |
| None | 23 | 3(13.0) |
| **Occupation** |  |  |
| Student | 25 | 10(40.0) |
| Non-student | 75 | 21(28.0) |
| **Religion** |  |  |
| Christianity | 96 | 30 (31.3) |
| Others | 4 | 1(25.0) |
| **Total** | **100** | **31(31.0)** |

### 3.2. Overall Prevalence of Syphilis, HCV, and *H. Pylori* among Pregnant Women

Of the 100 pregnant women tested for *Helicobacter pylori*, HCV, and syphilis at Mbodo Health Centre in Aluu, Rivers State, Nigeria, 31.0% were positive for *H. pylori* and none (0.0%) for HCV antibody and syphilis antigen (Table 1).

### 3.3. Prevalence of *H. Pylori* among Pregnant Women with their sociodemographic characteristics

According to the study, 26.7% of participants in the 20–29 age group tested positive for *Helicobacter pylori*. Of the 30-39 age group, 37.5% tested positive for *H. pylori* and the age range of 40-49 had the least prevalence (14.3%) as displayed in Table 1. A higher prevalence of *Helicobacter pylori* occurred among the married groups (32.7%) than in the unmarried group with a 29.4% prevalence. Forty-four per cent of the pregnant women who tested positive for *H. pylori* had attained secondary education. A higher prevalence of *Helicobacter pylori* occurred with secondary education than with tertiary education (32.7%) and no form of education (13.0%). Occupationally, a higher prevalence of *Helicobacter pylori* occurred among students (40.0%) than among non-students (28.0%). Based on religion, a higher prevalence of *Helicobacter pylori* occurred among Christians (31.3%) than among other religious groups (25.0%) as indicated in Table 1.

## 4. Discussions

Human infections such as syphilis, HCV, and *H. pylori* are dangerous to human populations. Pregnant women and their foetuses are particularly vulnerable to these infections. According to Gutwerk et al. (2018), they can spread congenitally or vertically to the foetus. The pregnant women in this study had zero prevalence of syphilis and HCV antibody.

Although syphilis control is still a difficult task (Arando et al., 2019), the overall prevalence in this study is 0.0%. This observation is consistent with the findings of Alubi et al. (2023) in Port Harcourt and with Okonko et al. (2012a, b, c, 2013) in Ibadan. However, the 0.0% observed in this study is lower than the 1.5% and 6.6% reported previously in Port Harcourt (Adewuyi-Oseni et al., 2019; Okonko et al., 2020a); 1.7% in Uyo (Okonko et al., 2020b) and in Enugu (Chukwurah & Nneli, 2005); the 2.0% and 2.63% prevalence observed in the metropolitan regions of Akwa Ibom State, Nigeria (Opone et al., 2020). Furthermore, the 0.0% syphilis report in this study is less than the 0.8% recorded in Ibadan, Nigeria by Okonko et al. (2012b). Furthermore, this frequency was lower than the 2.9% prevalence in sub-Saharan Africa (Hussen & Tadesse, 2019), the 0.3% observed in Eritrea (Siraj et al., 2018), and the 0.1% documented in Port Harcourt (Ejele et al., 2005).

As much as hepatitis C virus (HCV) infection is a serious global health concern, with estimates putting its prevalence between 2.0% and 3.0% of the global population (Okonko & Shaibu, 2023), the present study indicated a 0.0% HCV prevalence. Our observation is consistent with Okonko et al. (2014) and Cookey et al. (2021) who also reported a 0.0% prevalence in their studies. The 0.0% infection rate with HCV contradicts the 4.0% by Okonko and Shaibu (2023) in Yenogoa, Bayelsa State, Nigeria and Oketah et al. (2024) in Awka, Anambra State, Nigeria; the 15.0% and 23.5% reported by Ogbodo et al. (2015) and Ogwu-Richard et al. (2015) in Nigeria. This 0.0% is also lower than the 0.5% in Lagos (Lawal et al., 2020), the 1.0% was recorded by Aaron et al. (2021) in Port Harcourt, Nigeria, the 0.5% in Anyigba (Omatola et al., 2019), the 0.8% in Abuja (Agboghoroma & Ukaire, 2020), the 0.8% in Osogbo (Oluremi et al. (2021), the 22.5% in Port Harcourt, Nigeria (Okonko et al., 2022) and the 5.6% in Emohua, Rivers State, Nigeria (Okonko & Ernest Nwagwu, 2023).

*Helicobacter pylori* (*H. pylori*) is a gram-negative ubiquitous bacterium affecting over half of the world’s population (Emerenini et al., 2021). The seroprevalence of *H. pylori* among pregnant women in this study was 31.0%, which was less than the total burden of 70.17% and 87.7% in Nigeria and Africa, respectively that Smith et al. (2022a) reported. According to some academics, the reason behind the high prevalence of *Helicobacter pylori* is inadequate hygiene; as a result, half of the global population has *Helicobacter pylori* infection (Alexander et al., 2021). The majority of infected individuals (>80%) have asymptomatic chronic gastritis; only a tiny number of infected individuals eventually develop PUD or gastric malignancies (Smith et al., 2019a). Most developing nations have higher prevalence rates of *Helicobacter pylori*, which is typically correlated with socioeconomic level and hygiene (Hooi, 2017). Also, 2.6% of Nigerian patients had stomach cancer brought on by *H. pylori* (Smith et al., 2022b).

The results of the current study, which show 31.0%, conflict with those of Alubi et al. (2023) in a secondary health facility in Port Harcourt, Nigeria and some other areas in Nigeria and overseas with varying figures and proportions. Bello et al. (2018) reported 81.7% in Kano, Nigeria. Okoroiwu et al. (2022) reported 72.4% in Owerri, Imo State, Nigeria. Chen et al. (2014) reported 72.1% of *H. pylori* infection in Lanyu. Lawson (2023) reported 63.0% in Port Harcourt, Nigeria. Mnichil et al. (2023) reported 62.0% prevalence in Yilmana Densa District, Northwest Ethiopia. Ibrahim et al. (2022) observed a 58.9% prevalence in Gombe, Nigeria. Akhimienho et al. (2021) reported 45.7% in Port Harcourt, Nigeria. Okonko et al. (2016) and Okosigha (2014) reported 44.4%. Ahaotu et al. (2023b) observed a 38.0% prevalence in Port Harcourt, Nigeria.

The 31.0% reported here, however, is higher than the 2.0% observed in Port Harcourt, Nigeria (Okonko & Barine, 2023), the 12.7% reported by Jemikalajah and Okogun (2014) in Warri, Nigeria, the 19.6% by Ayodele et al. (2018) in Port Harcourt, Nigeria and the 20.0% reported by Ahaotu et al. (2023a) in Port Harcourt and Emerenini et al. (2021) in Owerri, both in Nigeria, the 26.2% reported by Innocent-Adiele et al. (2023) in Calabar, Nigeria and the 29.7% reported by Owowo et al. (2019) in Bakassi Peninsular and Etim Ekpo in South Southern, Nigeria.

In terms of age, women in the 30–39 age range had a greater *Helicobacter pylori* seroprevalence than the other age groups. In comparison, the study conducted by Hong et al. (2019) found that the majority of participants in the 30-34 age groups had a seroprevalence of *H. pylori* infection. This observation aligns with the findings of Zhu et al. (2014), who found that *H. pylori* infection is more common in 30-to 39-year-olds. Our observation is even with Ayodele et al. (2018) and Ibrahim et al. (2022), who found that N*H. pylori* infection is more common in the 31-to 40-year age group in Port Harcourt, and Lafia, respectively.

The present observation with age disagrees with that of Alubi et al. (2023) who reported a higher prevalence in 20-40 years age groups in Port Harcourt, Nigeria. Additionally, it contradicts the findings of Efere (2019) and Innocent-Adiele et al. (2023), who found a higher frequency in the 26–30 age range. Additionally, it contradicts the findings of Ahaotu et al. (2023a), who found that pregnant women in Port Harcourt between the ages of 20 and 29 had a greater frequency. This result also contradicts findings from studies conducted in Port Harcourt by Okonko and Barine (2023), Okosigha (2014), Okonko et al. (2016), and Okoroiwu et al. (2022) in Owerri, Imo State, Nigeria and Kooffreh-Ada et al. (2019) in Calabar, Nigeria, which demonstrated that *H. pylori* infection is rather common between the ages of 40 and 60. These studies also reported lower seropositivity in ages 0-18 and higher prevalence in 19-37. Owowo et al. (2019) reported the greatest prevalence among less than 10 years of age group in Bakassi Peninsular and Etim Ekpo in South Southern, Nigeria. Akhimienho et al. (2021) reported a higher prevalence in the age group 12-17 years in Port Harcourt, Nigeria. Kabido et al. (2022) indicated the highest prevalence among less than 15-year age groups in Lafia, Nigeria. Additionally, it contradicts the findings of Ahaotu et al. (2023b) and Lawson (2023), who found that *H. pylori*-infected individuals in Port Harcourt, Nigeria, were more likely to be in the 40–50 age range. Our observation is at odds with Mnichil et al. (2023) who reported the highest prevalence in the age group above 50 years in Yilmana Densa District, Northwest Ethiopia. This could happen because elderly people have been exposed to the pathogen for a longer period than younger people.

A higher prevalence of *Helicobacter pylori* occurred among the married groups than in the unmarried group, which is not consistent with Okonko and Barine et al. (2023) findings, which showed that singles had a higher prevalence. This can occur from the illness spreading through bodily fluids (by kissing saliva). However, the present observation is consistent with Okonko et al. (2016) and Innocent-Adiele et al. (2023) who reported a higher prevalence of *H. pylori* among the married in Port Harcourt and Calabar, Nigeria, respectively. Our observation is at even with Mnichil et al. (2023) who also reported the highest prevalence in the married groups in Yilmana Densa District, Northwest Ethiopia. Kabido et al. (2022) also indicated the highest prevalence among the married group in Lafia, Nigeria

People from lower socioeconomic classes are more likely to live in areas where there is a higher risk of faecal contamination of water and food, have lower levels of education, and receive poor health education (Bello et al., 2018; Alubi et al., 2023). At 44.0%, the seroprevalence was greater in those with secondary education than with post-secondary education (32.7%) and no form of education (13.0%). This is at odds with a study by Zhu et al. (2014) that showed illiterate persons had the greatest prevalence of *H. pylori* infection and university graduates had the lowest rates. It is also at odds with that of Okoroiwu et al. (2022) in Owerri, Imo State, Nigeria which showed that illiterate persons had the greatest prevalence of *H. pylori* infection. Our observation is at odds with Mnichil et al. (2023) who reported the highest prevalence among the illiterates in Yilmana Densa District, Northwest Ethiopia. Kabido et al. (2022) indicated the highest prevalence in those with informal education. A study by Owowo et al. (2019) showed that primary education holders had the greatest prevalence. This could be because those with higher levels of education may be more exposed and, thus, have had more interaction with the condition. According to Abdollahi et al. (2014), there was no discernible difference in the distribution of *H. pylori* between the two groups despite their attempts to match the cases and controls for residency and educational attainment.

Bello et al. (2018) found that being from a lower socioeconomic class increases your risk of getting an *H. pylori* infection. Occupationally, a higher prevalence of *Helicobacter pylori* occurred among students (40.0%) than among non-students (28.0%). The present observation agrees favourably with the findings of the study by Efere (2019) and Innocent-Adiele et al. (2023), who designated a higher prevalence among teachers, followed by students in Calabar, Nigeria. According to research by Ishaleku and Ihiabe (2010), the frequency of *H. pylori* infection rose from 45.5% in students to 85.7% in adults. This finding by Ishaleku and Ihiabe (2010) was not consistent with the higher seroprevalence rate of 40.0% for students in the employment category. The present study is consistent with that of Alubi et al. (2023) and Okoroiwu et al. (2022) who reported more *Helicobacter pylori* infections among the unemployed groups (students inclusive). The present observation backs up the claim made by Smith et al. (2018) that a person’s profession has nothing to do with their *H. pylori* infection. The current results also contradict the findings of Ahaotu et al. (2023b), who asserted a larger prevalence of traders. It also contradicts the findings of Owowo et al. (2019), Kabido et al. (2022) and Mnichil et al. (2023), who asserted a larger prevalence of farmers.

Based on religion, a higher prevalence of *Helicobacter pylori* occurred among Christians. This could be partly due to the larger proportion of Christians enrolled in this study. All of these could be the result of people in prevalence categories with greater levels of carelessness and poor cleanliness, which leaves them vulnerable to *Helicobacter pylori*.

## 5. Conclusion

The findings of this study indicated that the three pathogens under investigation did not co-infect. This study’s findings demonstrated that the population is free of both HCV and syphilis. However, a high proportion of the pregnant women had *H. pylori*, indicating endemicity in the study area. It was found that pregnant women in their middle years were more likely to contract *H. pylori*. The current study’s findings serve as a baseline for further research into *H. pylori* infection in the South-South region of Nigeria. They represent the second documented prevalence of *H. pylori* among pregnant women in Port Harcourt, Nigeria by our team. To properly evaluate the infection rate among these pregnant women in Rivers State, Nigeria, more investigation is necessary.

## Data Availability

All data produced in the present work are contained in the manuscript.

## Compliance with ethical standards

## Acknowledgement

The authors appreciate everyone who agreed to participate in the study and the administration of the Primary Health Centre, Mbodo-Aluu, Rivers State, Nigeria, for their approval.

## Statement of ethical approval

All authors declare that all experiments have been examined and approved by the University of Port Harcourt Research Ethics Committee and University of Port Harcourt Teaching Hospital Research Ethics Committee. Therefore, the study is performed following the ethical standards laid down in the 1964 Declaration of Helsinki.

## Statement of informed consent

“All authors declare that informed consent was obtained from all individual participants included in the study.”

